# Surveillance of seasonal respiratory viruses among Chilean patients during the COVID-19 pandemic

**DOI:** 10.1101/2021.07.16.21260648

**Authors:** Luis A. Alonso-Palomares, C. Joaquín Cáceres, Rodrigo Tapia, Paulina Aguilera-Cortés, Santiago Valenzuela, Fernando Valiente-Echeverría, Ricardo Soto-Rifo, Aldo Gaggero, Gonzalo P. Barriga

**Author notes:** To whom correspondence should be addressed: Dr. Gonzalo Barriga, Emerging Viruses Laboratory, Virology Program, Institute of Biomedical Sciences, Faculty of Medicine, Universidad de Chile, Independencia 1027, Santiago, Chile.

## Abstract

SARS-CoV-2 has generated over 122 million cases worldwide. Non-pharmaceuticals interventions such as confinements and lockdowns started in Chile on March 18^th^ 2020. In Europe, confinements and lockdowns have been accompanied by a decrease in the circulation of other respiratory viruses such as Influenza A virus(IAV), Influenza B virus(IBV) or respiratory syncytial virus(RSV) (1). Although changes in circulation patterns of respiratory viruses have been reported, limited information regarding the southern hemisphere is available where the SARS-CoV-2 pandemic merged with the winter season. We conducted viral surveillance of respiratory viruses and we evaluated their presence and establishing whether they were co-circulating with SARS-CoV-2.

---

Few south hemisphere countries reported the same pattern than Europe where non-pharmaceutical measures began before the winter season (2, 3) but to the best to our knowledge, no report has been generated containing information from Chile. Here, we collected 800 nasopharyngeal-swabs samples (NSS) from 13 health care centers belonging to the north area of Santiago, Chile, between April 1^st^ to July 31^st^, 2020 (Figure 1). All samples were collected from patients with at least one COVID-19 symptoms. 400 samples were determined as positive for SARS-CoV-2. 64% percent of SARS-CoV-2 positive individuals showed age range between 23-57 years. In addition, women had a significant incidence of positive cases, corresponding to 59% in age range group (Figure 2).

**Figure 1:**
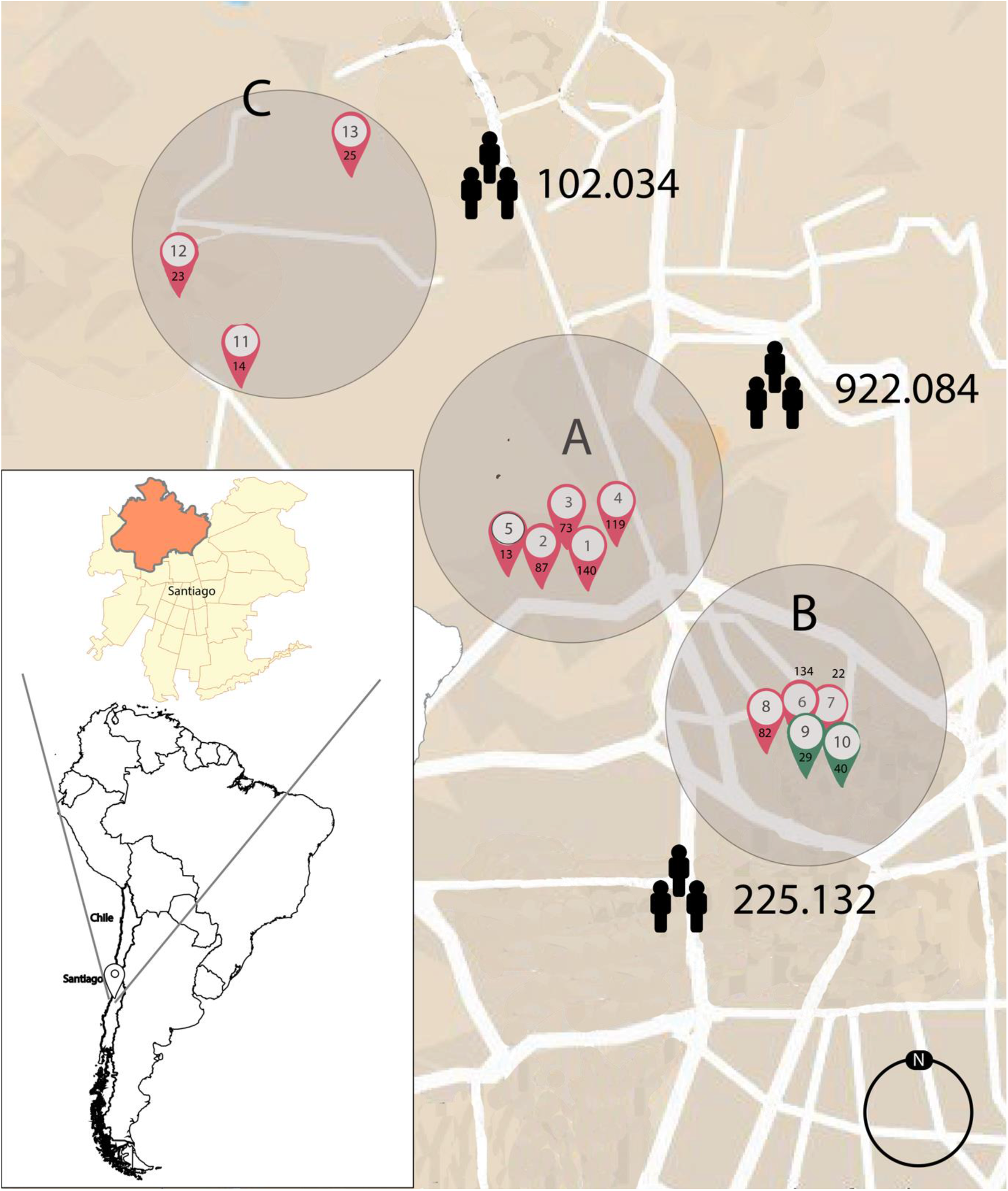
Geographical distribution of the samples analyzed in this study. Distribution of the thirteen-health care center from where the samples were obtained. The figure shows the number of samples by health care center (black number below white circle). Red flag shows health cares center with SARS-CoV-2 positive cases, green flag shows health cares with SARS-CoV-2 negative cases, the human shape indicate population by location (A, B and C).

**Figure 2.**
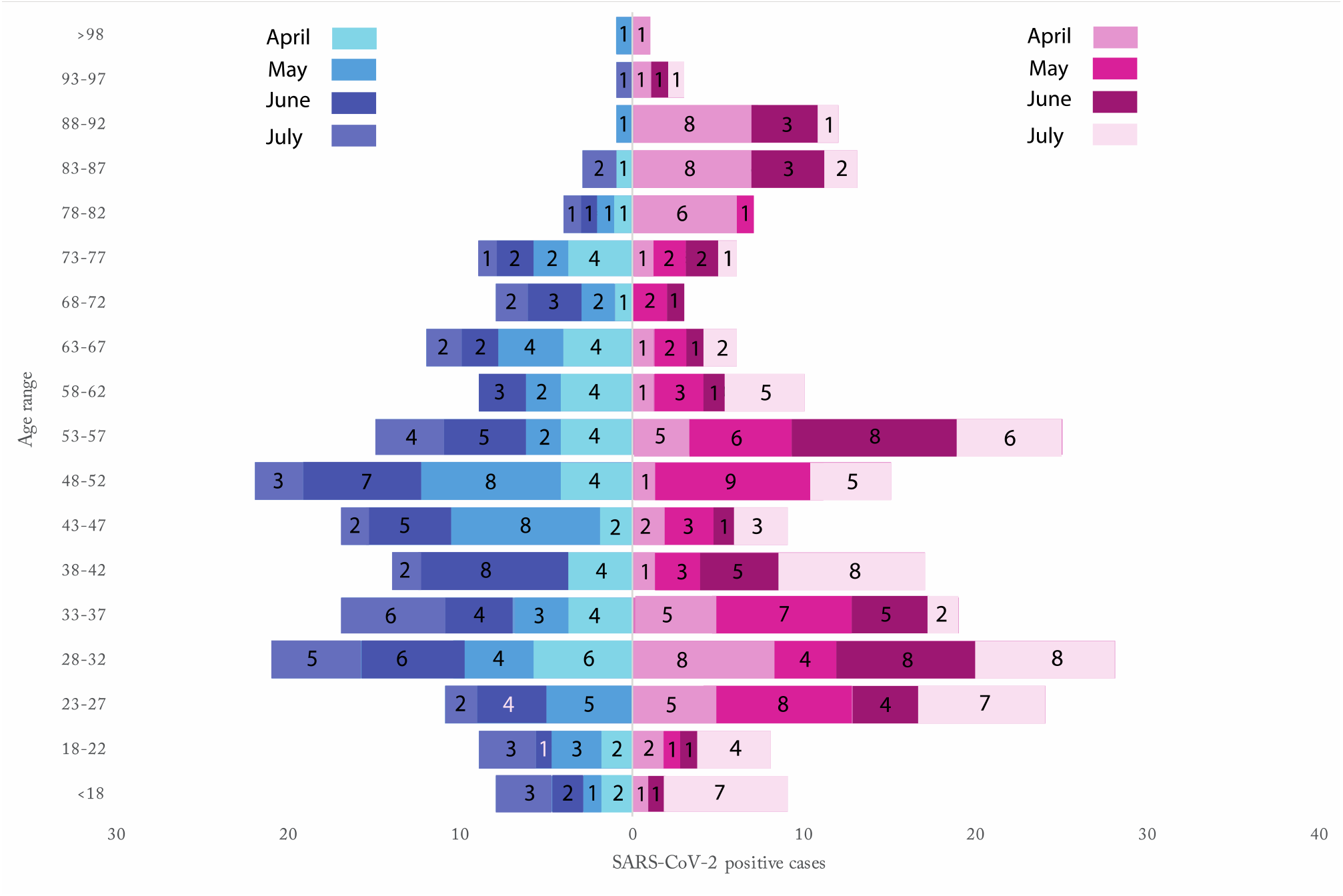
Number distribution of COVID-19 cases according to age group and sex. from April 1^st^ to July 31^st^ (2020) at Santiago of Chile. 400 patients are considered positive for SARS-CoV-2; however, three patients did not provide any information about age and gender. In red is shows female gender and in blue is shows male gender. The numbers in the columns indicate SARS-CoV-2 positive cases by month.

Next, based on geographic location we divided the samples in 3 groups (A, B and C) Figure 1. Location (A) contains the highest number of SARS-CoV-2 cases, contributing more than 50% of the positive samples analyzed in this study (Figure 1). This high positivity could be explained by the population density of location A, which contains at least 4-fold more inhabitants than locations B and C (Figure 1). Nevertheless, the health centers located in B presented positivity rates higher than 61% for SARS-CoV2 (Table 1), except for sub-locations 9 and 10 (Figure 1), where no positive samples were obtained in the period analyzed. Location C is farthest location form downtown, however, still has a positivity of 54.8% indicating a homogeneous distribution of SARS-CoV-2.

**Table 1.**
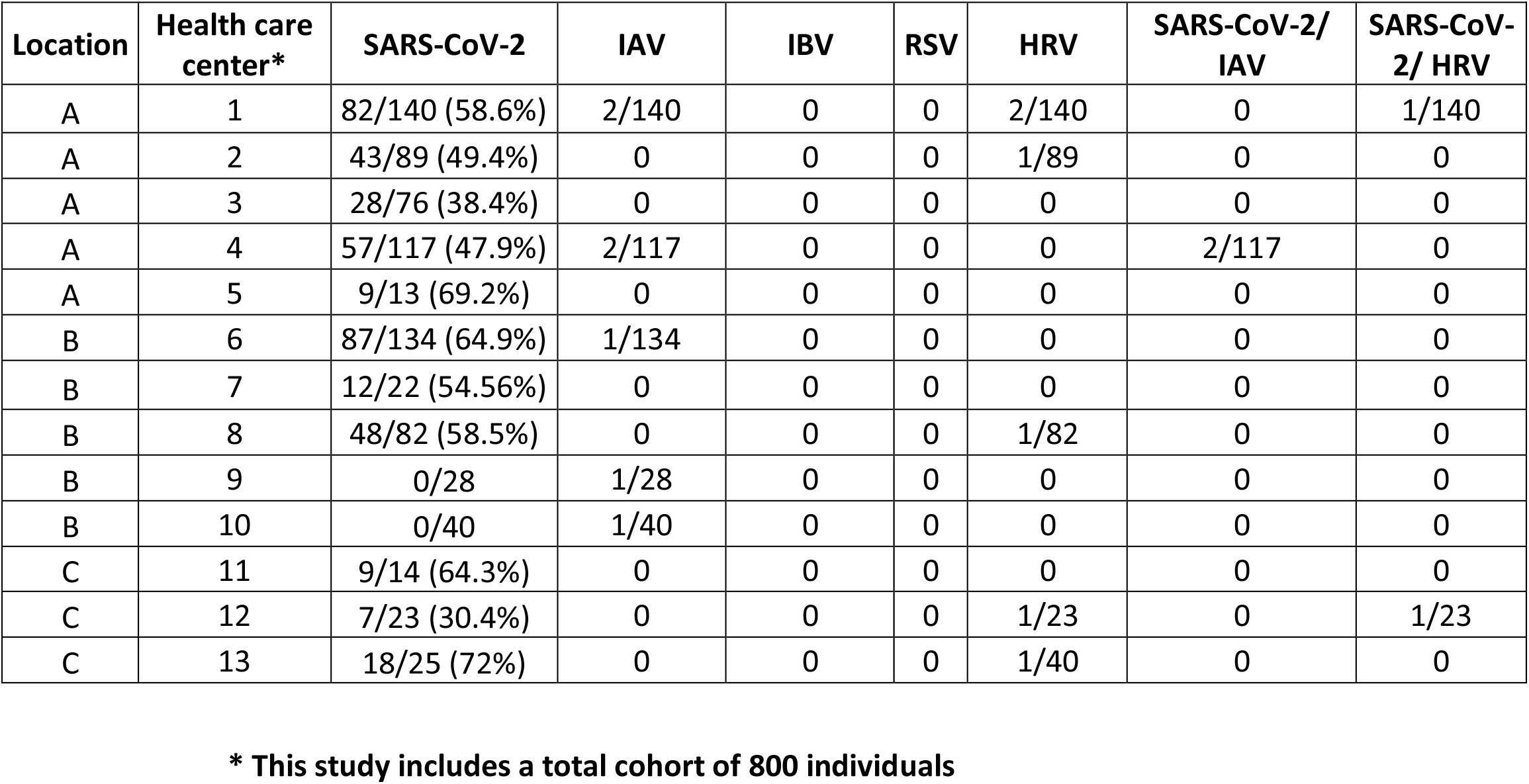
Co-circulation and co-infection of seasonal respiratory viruses together with SARS-CoV-2 in the Northern area of Santiago, Chile.

| Location | Health care center* | SARS-CoV-2 | IAV | IBV | RSV | HRV | SARS-CoV-2/<br>IAV | SARS-CoV-2/<br>HRV |
| --- | --- | --- | --- | --- | --- | --- | --- | --- |
| A | 1 | 82/140 (58.6%) | 2/140 | 0 | 0 | 2/140 | 0 | 1/140 |
| A | 2 | 43/89 (49.4%) | 0 | 0 | 0 | 1/89 | 0 | 0 |
| A | 3 | 28/76 (38.4%) | 0 | 0 | 0 | 0 | 0 | 0 |
| A | 4 | 57/117 (47.9%) | 2/117 | 0 | 0 | 0 | 2/117 | 0 |
| A | 5 | 9/13 (69.2%) | 0 | 0 | 0 | 0 | 0 | 0 |
| B | 6 | 87/134 (64.9%) | 1/134 | 0 | 0 | 0 | 0 | 0 |
| B | 7 | 12/22 (54.56%) | 0 | 0 | 0 | 0 | 0 | 0 |
| B | 8 | 48/82 (58.5%) | 0 | 0 | 0 | 1/82 | 0 | 0 |
| B | 9 | 0/28 | 1/28 | 0 | 0 | 0 | 0 | 0 |
| B | 10 | 0/40 | 1/40 | 0 | 0 | 0 | 0 | 0 |
| C | 11 | 9/14 (64.3%) | 0 | 0 | 0 | 0 | 0 | 0 |
| C | 12 | 7/23 (30.4%) | 0 | 0 | 0 | 1/23 | 0 | 1/23 |
| C | 13 | 18/25 (72%) | 0 | 0 | 0 | 1/40 | 0 | 0 |

**\* This study includes a total cohort of 800 individuals**

Taken together, our data show a high frequency of positive samples throughout the healthcare centers evaluated suggesting the population density as a risk factor for SARS-CoV-2 transmission since location A and B concentrates more population than location C. These results demonstrate that the 2020 winter season in Santiago presented a high incidence of SARS-CoV-2.

Then, we sought to determine in all samples whether SARS-CoV-2 was co-circulating with other respiratory viruses. We chose predominant respiratory viruses in Santiago (winter season), such as: IAV, IBV, RSV and human rhinovirus (HRV) (Table 1). Adenovirus, Parainfluenza and Metapneumovirus were not evaluated since they are considered all-year viruses (4). The results showed three samples with co-infection between IAV and SARS-CoV-2 (Table 1). This is congruent to recent studies in Ecuador and brazil showing complete decrease of IAV (5, 6). Despite the co-circulation or co-infection between IAV and SARS-CoV-2 observed, we could not detect RSV or IBV in the SARS-CoV-2 positive samples. These results suggest an impact of the non-pharmaceutical interventions in the circulation of seasonal viruses, as previously reported in Korea and Hong Kong (7, 8). Next, we evaluated the presence of IAV, IBV and RSV in the samples reported as negative for SARS-CoV-2 where five positive samples for IAV and no positive samples for IBV or RSV were detected, suggesting a circulation of these viruses below 1% considering the amount of samples evaluated (Table 1). This is lower than the information from the northern hemisphere where a range between 2-10% has been reported (9, 10). Taken together, these results suggest a lower co-circulation of IAV with SARS-CoV-2 and co-circulation below the level of detection of this study for SARS-CoV-2 together with IBV or RSV.

Finally, we focused on HRV, responsible for more than 50% of the cold-like illnesses, with a high preponderance to coinfection with other respiratory viral pathogens(11). Furthermore, HRV was the predominant virus after SARS-CoV-2 detected either cocirculating with SARS-CoV-2 or circulating alone(9). The presence of HRV was assessed, showing that 0.25% of the samples were co-infected SARS-CoV-2/HRV. On the other hand, the HRV co-circulation was 0.8% (Table 1). These results establish HRV co-circulation and the co-infection with SARS-CoV-2. Taken together, these results demonstrate the displacement of seasonal respiratory viruses due to the presence of SARS-CoV-2. Despite of this displacement, IAV and HRV are still able to keep cocirculating together with SARS-CoV-2 but to a considerably lesser extent in comparison with previous winter seasons.

## Discussion

To gain insights into the potential co-circulation of the most relevant seasonally circulating respiratory viruses together with SARS-CoV-2, a fact on going COVID-19 pandemic was that the vast majority of the SARS-CoV-2 testing during the April-July period was indicated only with the presence of symptoms, we arbitrarily selected 200 samples per month (April to July) for a total of 800 NSS from 13 health care centers located in the north zone of Santiago, Chile.

We detected a high positivity rate by health care center between 30,4%-72% and we observed at least twice co-infections between SARS-CoV-2/IAV or SARS-CoV-2/HRV and no co-infections with IBV and RSV, which is in agreement with previously reported data including the southern hemisphere (5,12). Furthermore, IAV and HRV were detected from negative SARS-CoV-2 samples, whereas no presence of IBV or RSV was obtained even from the negative SARS-CoV-2 samples (Table 1). These results demonstrate the displacement of the predominant seasonal respiratory viruses, which have an essential impact during the winter season caused by the high circulation rate of SARS-CoV-2. A similar phenomenon was observed after the 2009 Influenza A (H1N1) pandemic, which generated a decrease of RSV and IAV H3N2 infections (13). The reduction or absence of IAV, IBV or RSV observed in this study can be explained by the non-pharmaceutical interventions such as confinement and lockdowns established before the beginning of the winter season in March 2020. A previous report showed that SARS-CoV-2 could replace within three weeks the seasonal respiratory viruses circulating (1), while that HRV co-infections are one of the most commonly observed. However, the impact that HRV infection co-infecting with other respiratory viruses is still unclear due to inconsistencies among different studies (11). The effect of HRV in SARS-CoV-2 infection and the clinical outcome is still unknown.

Considering that the vast majority of the SARS-CoV-2 testing during the April-July period was indicated only with the presence of symptoms, potential bacterial infections or co-infections cannot be ruled out in this study. The presence of bacterial infections during the SARS-CoV-2 pandemic has been previously reported (14-16). A previous study identified *S. Pneumoniae, K. pneumoniae and H. influenza* among the bacteria cocirculating with SARS-CoV-2 (16). However, the detection of bacteria is beyond the scope of the study In conclusion, the data shows the impact of SARS-CoV-2 over the co-circulation of seasonal respiratory viruses like IAV, IBV, and RSV in Chile. Our results suggest that the emergence of SARS-CoV-2 in addition with different non-pharmaceutical measures adopted worldwide have a detrimental impact on the circulation at least of seasonal respiratory viruses. Furthermore, our data allow us to foresee the circulation of respiratory viruses in the 2021 winter season in the southern hemisphere.

## Data Availability

The data that support the findings of this study are available upon request from the authors

## Ethical statement

The study described here was approved by the Ethics Committee of the Faculty of Medicine at Universidad de Chile (Project Nº 036-2020). The samples were de-identified and not considered as human samples.

## Acknowledgments and Funding

The authors are supported by Instituto Antártico Chileno (INACH) RT_35-19 (GB-P), ANID Chile through Fondecyt grants Nº 11200228 (GB) 1181656 (AG), 1190156 (RS-R), 1180798 (FV-E); Postdoctoral fellowship Nº SECTEI/138/2019 from Mexico City (LA-P). Authors would like to thank the Science, Technology, Knowledge and Innovation Ministry of Chile for articulating and coordinating support from the scientific community. Also, we want to thank the diagnostic group of the University of Chile

## Authors contributions

Conceptualization: LAP, CJC and GB, Data curation: LAP, RT, PA, AG, FV-E and GB, Formal analysis: LAP and GB, Funding acquisition: GB, Investigation: LAP, RT, PA, SV and GB, Methodology: LAP, CJC, FAV-E, AG, RS-R and GB, Project administration: LAP, CJC, FV-E, AG, RS-R and GB.

Supervision: LAP, FAV-E, AG, RS-R and GB, Validation: LAP, CJC, and GB, Visualization: LAP, CJC, and GB, Writing-original & draft: CJC, and GB, Writing-review & editing: LAP, CJC, FAV-E, AG, RS-R and GB, All authors approved the final version of the manuscript. Gonzalo Barriga had full data access to all data in this study and takes complete responsibility for the integrity of the data and the accuracy of the data analysis.

## Transparency statement

Gonzalo Barriga affirms that this manuscript is an honest, accurate, and transparent account of the study being reported; that no important aspects of the study have been omitted; and that any discrepancies from the study as planned have been explained

## Data availability statement

The authors confirm that the data supporting the findings of this study and its supplementary materials.

## Materials and methods

### Sample selection

All samples in this study were obtained with nasopharyngeal swabs come from suspected Chilean population infected with SARS-CoV-2 in the north of Santiago de Chile from different health centers (figure 1). The samples were collected in 2 mL of RNA-shield media (GenoSUR) and store at room temperature until its analysis for SARS-CoV-2 detection.

### RNA extraction and Identification of respiratory viruses by RT-qPCR

The RNA extraction was made using the Total RNA Purification Kit (Norgen Biotek CORP); following the manufacturing procedure, the RNA was a store at -80°C, which was used to perform RT-qPCR.

All samples were analyzed using a specific primer (Supplementary table 1) for SARS-CoV-2, IAV, IBV, RSV, and HRV. All sequences have been validated, and their use is a typical procedure to detect respiratory viruses from the World Health Organization (WHO). The viral genome detection was made using LightCycler® Multiplex RNA Virus Master (Roche) following the manufacturing procedure. The amplification and analysis plot was made in a QuantStudio3 Real Time PCR System 96 wells (Thermo Fisher Scientific). The HRV and IBV detection was performed by RT-PCR final point using specific primers, the retrotranscription step was made using SuperScript IV Reverse Transcriptase (Thermo Fisher Scientifics) and PCR was using GoTaq® DNA polymerase (Promega), the genome of HRV was visualized in agarose – Seakem LE Agarose (LONZA) at 2% applying a voltage of 80 Volts for 30 minutes.

**Supplementary Table 1.**
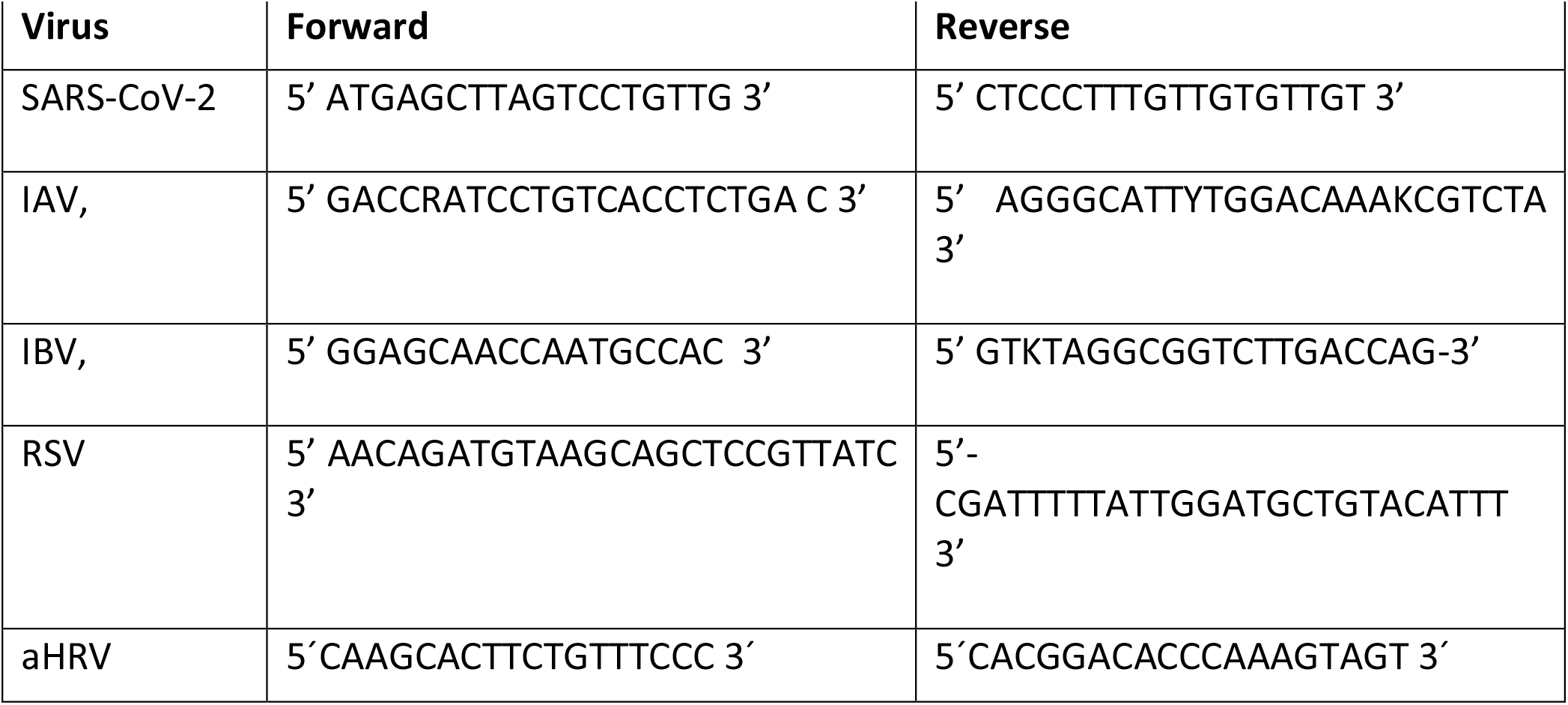
Primers Sequence for SARS-CoV-2, IAV, IBV, RSV and HRV.

